# Burden and correlates of complicated severe acute malnutrition relapse among children under five at Mwanamugimu: *Secondary analysis of hospital data in Uganda*

**DOI:** 10.1101/2025.10.01.25337123

**Authors:** Joseph Mbabazi

## Abstract

**Background:** Confronted with the triple burden of malnutrition including in low- and middle-income country settings (LMICs), effort has been made to tackle child wasting especially to improve treatment outcomes of complicated severe acute malnutrition (cSAM). However, some children relapse after recovery and are retaken through the elaborate management criteria which not only strains the system, but also puts a toll on their health.

**Objective:** We assessed the proportion of cSAM relapses and the associated socioeconomic, anthropometric, and clinical factors.

**Method:** This was a comparative cross-sectional study using secondary data of children under 5 with cSAM admitted at Mwanamugimu nutrition unit between May 2015 to April 2017. Multiple logistic regression analysis was used to assess for the correlates of cSAM.

**Results:** We included 1098 children with mean age 13.8 ±9.8 months, over two-thirds were male, and ∼50% hailed from Kampala. On analysis, 5.7% were cSAM relapses, 0.7% were multiple relapses, and it took them ∼36 weeks on average to relapse. Increased age, number of siblings, and children with unemployed mothers had 0.05 (95%CI: 0.02; 0.08), 0.3 (95%CI: 0.09; 0.51), and 2.63 (95%CI: 0.61; 4.65) higher odds of relapsing with cSAM respectively. There was no observed association with any anthropometric or clinical factor.

**Conclusion:** Despite lack of a standard definition and set threshold, a compelling proportion of children relapse with cSAM seemingly unnoticed across CMAM programs in LMICs but this may be at a cost. Treatment programs need to ensure they establish readmissions and also reinforce their community leakages so that poor, large households are benefactors of livelihood support to cater for underlying factors and mitigate risk of cSAM relapse.

## Introduction

Malnutrition, especially in the wake of its triple burden, is still a major public health challenge affecting mostly children in Africa and Asia [1]. Over 45 million children under 5 are affected by wasting which is a life-threatening condition mainly as a result of poor nutrient intake and/or recurrent illnesses [1, 2]. In 2022, 70% of all children affected by wasting lived in Asia and >¼ lived in Africa with 1 in every 5 in Sub-Saharan Africa facing hunger partly due to ongoing conflicts, drought, surging food prices, inequity, and weak infrastructural development [3]. Severe child wasting (severe acute malnutrition [SAM]) is defined as a weight-for-height z-score (WHZ) < -3 or a mid-upper arm circumference (MUAC) < 11.5 cm [4] and both are predictors of increased mortality MUAC being more sensitive [5]. Additionally, presence of bilateral pitting oedema is also classified as SAM. In Uganda, although the proportion of children with wasting has declined gradually to 3.2%, there is a plodding increase in child overweight/obesity (3.4%) on top of micronutrient deficiencies including nutritional anaemia (53.0%) [6]. This highlights the emerging trend even in such low- and middle-income country settings (LMICs) moreover, emerging data shows a tendency for stunting to develop concomitantly with wasting with increased mortality risk [7]. Addressing undernutrition has been key in the country with implementation of the integrated management of acute malnutrition (IMAM) strategy at the fore [8]. Here, all complicated SAM (cSAM) children defined by having SAM with medical complications, are managed in inpatient therapeutic care (ITC) [8]. Thereafter, children are discharged through the outpatient therapeutic care (OTC) where, together with uncomplicated SAM cases, they are managed using home-based ready-to-use therapeutic food (RUTF) alongside other services. Mwanamugimu nutrition unit (MNU) at Mulago national referral and teaching hospital is a center of excellence offering both ITC and OTC on top of other services. Although the strategy has achieved some milestones such as reduced length of hospital stay, notable challenges persist including relapses which not only pose a great strain on the system but also endanger children’s lives. In a previous systematic review, 0 – 37% children relapsed after SAM treatment but with varying lengths of time post-discharge and orphans had 5 times greater odds of relapse six months post-discharge [9] Despite lack of a clear definition and set threshold for cSAM relapse both in national guidelines [8] and global SPHERE standards [10], the crude annual 5% relapse of ∼1200 patients/year admitted between 2006 – 2014 in MNU clinical reports is unscientifically studied. Moreover, the factors associated with these relapses are unclear in spite of the elaborate management criteria. Therefore, we sought to assess the prevalence and associated socioeconomic, nutritional, plus clinical factors of cSAM relapses. Our results will not only reveal the proportion of affected children but also inform programmers, especially in LMICs, on factors to observe for effective care outcomes.

## Methods

### Study design

This was a comparative cross-sectional study using retrospective data from children under 5 years managed for cSAM at Mwanamugimu between May 2015 – April 2017. From May 1^st^ 2017 and guided by the unit’s medical records staff, we gained access to the stacked hard copy files retrieving a monthly stack at a time and later re-stacked by the records officer. During data collection, the research assistants and principal investigator had access to patient identify information but not thereafter as these were all coded with unique IDs after abstraction. The current data contributed to my dissertation for my Masters of Public Health Nutrition.

### Study setting and participants

The study was conducted at Mwanamugimu nutrition unit which is part of the paediatric units of Mulago national referral and teaching hospital in the capital city of Kampala, Uganda. Starting with April 2017, we consecutively sampled out files of children under 5, previously discharged from the unit and in case of readmissions, we checked out for their previous file which gave the index admission data. We excluded children who passed on, ran away, or were aged above 5 years.

During ITC management for cSAM, children were taken through three stages of stabilization, transition, and rehabilitation. In stabilization, they were covered with a broad-spectrum antibiotic intravenously while being fed on the starter therapeutic milk Formula 75 (F75) for 3 – 5 days. Having stabilized, they were given an acceptance test for RUTF and then gradually transitioned to this while reducing on F75 quantity until eventually being promoted to rehabilitation. In rehabilitation, as part of their diet, children were offered high energy porridge (consisting of corn, soy, milk [JESA]) and *kitobeero* a nutritious therapy made using locally available foods offering ∼850 kcal/day (**Table 1**). After ∼21 days of ITC management, children were discharged through OTC where they returned fortnightly for medical follow-up and RUTF refill until full recovery from SAM.

**Table 1:** Estimates of energy contents and macronutrient composition of drinks/foods served at Mwanamugimu.

| Feed: | Kcal/100 ml | Protein/100 ml |  | Fat/100 ml |  | Carbohydrate/100 ml |  | Vitamin/Mineral<br>(yes, no) |
| --- | --- | --- | --- | --- | --- | --- | --- | --- |
|  | Kcal | g | E% | g | E% | g | E% |  |
| Formula 75<sup>1</sup> | 76 | 0.9 | 4.7 % | 2.6 | 30.7% | 12.2 | 64.6% | yes |
| Formula 100<sup>2</sup> | 101 |  | >10% |  | >45% |  | <45% | yes |
| CornSoyaBlend<sup>3</sup> | 140.3 | 4.4 | 13% | 5.5 | 35% | 18.3 | 52% |  |
| Kitobeero<sup>4</sup> |  |  |  |  |  |  |  |  |
| with beans: | Total: 837 kcal |  | 19% |  | 6% |  | 75% |  |
| with groundnut: | Total: 889 kcal |  | 13% |  | 52% |  | 35% |  |
| Yoghurt<sup>5</sup> | 61 | 3.3 | 21.5% | 3.3 | 48.5% | 4.6 g | 30% |  |
| ReSoMal | 14 | - | - | - | - | 3.5 g |  | Na, K, Mg, Zn, Cu |
| RUTF | 520 | 12.8 | 10% | 30.2 | 45% | 41 |  |  |

<sup>1</sup>Serial number: 27517 L1112; <sup>2</sup>Serial number: L2096; <sup>3</sup>Made using 25 kg corn-soya flour (2:1 corn:soya), 3 litres refined vegetable cooking oil fortified with vitamin A, 3 kg sugar; <sup>4</sup>Made using 1 palm carbohydrate (corn flour or rice), 2 palms plant protein (beans or groundnuts), and 1 teaspoon of silver fish powder; <sup>5</sup>Made using 100 ml JESA full cream milk inoculated with culture from previous yoghurt
**Note:** 1 g carbohydrate – 4 kcal; 1 g protein – 4 kcal; 1 g fat – 9 kcal. Macronutrient calculation based on foodcomp.dk
Adopted from FeedSAM study [11] data collected by Charlotte Gylling Mortensen and Joseph Mbabazi

### Outcomes

A cSAM relapse defined as any child under 5 years readmitted to ITC ≥3 months post-discharge. Despite lack of an already established national or global definition, this was based partly on the fact that the expected average length of stay on OTC is set at 3 months [8, 10]. Children had been readmitted with SAM plus at least a medical complication or IMCI danger sign per guideline. The nine associated medical complications included; hypoglycaemia, hypothermia, severe infections like pneumonia, severe dehydration, shock, severe anaemia, cardiac failure, severe dermatosis, and corneal ulceration. Conversely, the IMCI danger signs were; poor appetite, intractable vomiting, convulsions, lethargy, unconsciousness, inability to breastfeed/drink, high grade fever, grade-3 oedema, and failed appetite test.

### Potential correlates of cSAM relapse

A data extraction tool was used to capture data on child’s sociodemographic and socioeconomic data such as age, sex, religion, residence, and caregiver’s occupation among others. For occupation, caregivers were considered employed if indication was made of being engaged in any formal or informal income-generating activity such as doing laundry around their neighborhoods.

Data was also captured regarding children’s nutrition status including presence of nutritional oedema, admission weight, height, and MUAC. The respective z-scores were generated in emergency nutrition assessment for standardized monitoring and assessment of relief and transitions (ENA for SMART, version 2011). The WHO cutoff points [4] were used to calculate WHZ, WAZ, and HAZ and all data exported and analyzed in StataSE 13.0 Clinical data on children’s HIV status, immunization, presenting medical complications or IMCI danger signs, and laboratory investigations were also noted.

### Data management and statistical analyses

Data was collected using electronic structured questionnaires in open data kit (ODK) platform with inbuilt checks. This allowed use of skip logic and minimized entry of any ambiguous data. Descriptive statistics were reported as %(n), and mean ±SD. Logistic regression analysis was done to assess for potential correlates of cSAM relapse at 95% confidence interval and 5% significance level. We minimally adjusted only for the universal confounders age and sex to minimize on introducing any potential bias. At 80% power, an alpha level of 5%, with 5% proportion of relapses among HIV negative children, and an odds ratio of 2, we needed 517 children per group and 1034 children in total. After data retrieval, we included 1098 in our analysis

### Ethical statement

Study approval was obtained from Makerere University School of Public Health Higher Degrees Research and Ethics Committee. Joint approval was obtained by the school from Uganda National Council of Science and Technology for all the Master students. Separate approval was sought from the Mulago hospital ethics and research committee (MREC: 1153) which was used to seek further permission from the hospital director and head of Mwanamugimu nutrition unit.

## Results

### Prevalence of cSAM relapse

Data from 1098 children under five admitted at MNU from May 2015 to April 2017 was reviewed. Nearly 6% of the children admitted during this time were cSAM relapses (**Figure 1**). Three children out of those that relapsed (representing 4.8%) were multiple relapses with cSAM (relapsed more than once) and it took 251 days (∼36 weeks) on average for a child to relapse.

**Figure 1:**
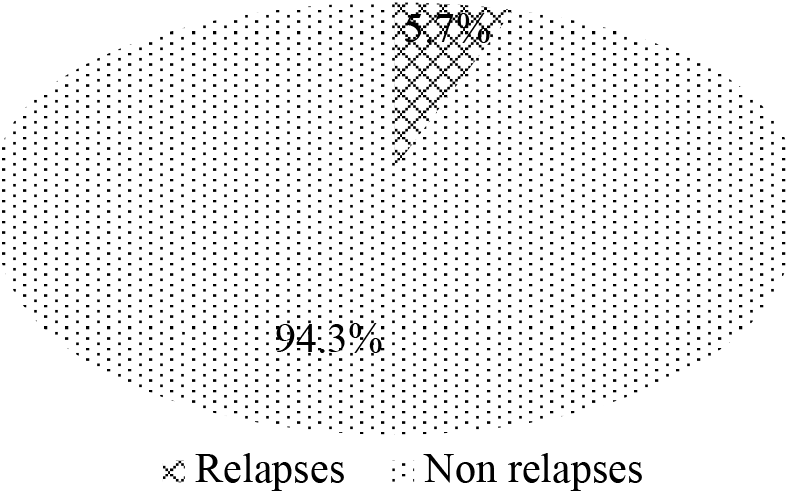
Prevalence of complicated SAM relapse at Mwanamugimu, Mulago hospital

### Correlates of cSAM relapse

Children were aged 13.8 ±9.8 months on average at their index admission with majority under two years of age that is 85.7% and 85.8% among relapses and non-relapses respectively (**Table 2**). Over two-thirds were male and nearly half hailed from Kampala district (the capital) without major differences in proportion between relapses (46.0%) and non-relapses (43.5%) (Table 2). Over 1/6^th^ were admitted with secondary caregivers (15.9% vs 15.0% between relapses vs non-relapses) and slightly over half of the caregivers in both groups were married that is 54.0% among relapses vs 56.8% among non-relapses (Table 2).

**Table 2:** Baseline characteristics of 1098 children with complicated severe acute malnutrition admitted for inpatient therapeutic care at Mwanamugimu, Mulago hospital between May 2015 – April 2017^1^.

| Variables |  | *n* | Relapses<br>*n*=63 | Non relapses<br>*n*=1035 |
| --- | --- | --- | --- | --- |
| **Sociodemographic and socioeconomic data** |  |  |  |  |
| Age (months) | <24 | 1098 | 85.7% (54) | 85.8% (888) |
|  | 24 – 59 |  | 14.3% (9) | 14.2% (147) |
| Sex | Male | 1094 | 63.5% (40) | 60.3% (624) |
| Residence | Kampala | 1096 | 46.0% (29) | 43.5% (450) |
|  | Wakiso |  | 23.8% (15) | 35.1% (363) |
|  | Other districts |  | 30.2% (19) | 21.5% (222) |
| Caregiver – child relationship | Mother | 1098 | 84.1% (53) | 85.0% (880) |
|  | Caretaker |  | 15.9% (10) | 15.0% (155) |
| Number of surviving children at home |  | 920 | 3.1 ± 1.8 | 2.6 ± 1.5 |
| Caregiver's marital status | Married | 1098 | 54.0% (34) | 56.8% (588) |
|  | Not married |  | 46.0% (29) | 43.2% (447) |
| Caregiver's occupation | Employed | 1098 | 19.1% (12) | 30.9% (320) |
|  | Unemployed |  | 60.3% (38) | 43.2% (447) |
|  | Missing data |  | 20.6% (13) | 25.9% (268) |
| **Nutritional data** |  |  |  |  |
| Nutrition diagnosis | Severe acute malnutrition non-oedematous | 1098 | 79.4% (50) | 53.4% (553) |
|  | Severe acute malnutrition oedematous |  | 17.5% (11) | 40.4% (418) |
|  | Moderate acute malnutrition |  | 3.2% (2) | 6.2% (64) |
| Weight (kg) |  | 1046 | 6.8 ± 2.1 | 6.0 ± 2.9 |
| Mid-upper arm circumference (cm) |  | 278 | 11.4 ± 2.1 | 10.7 ± 1.2 |
| Weight-for-age z-score |  | 636 | -4.2 ± 0.8 | -4.4 ± 1.5 |
| Height-for-age z-score |  | 955 | -3.0 ± 1.7 | -2.9 ± 2.1 |
| Weight-for-height z-score<sup>2</sup> |  | 589 | -4.4 ± 1.1 | -3.8 ± 1.6 |
| **Clinical data** |  |  |  |  |
| HIV status | HIV sero-exposed | 1098 | 11.1% (7) | 20.5% (212) |
|  | Negative |  | 22.2% (14) | 59.8% (619) |
|  | Unknown |  | 9.5% (6) | 17.1% (177) |
|  | Missing data |  | 57.1% (36) | 3.0% (31) |
| Immunization status | Up-to-date<sup>3</sup> | 1098 | 15.9% (10) | 40.0% (414) |
|  | Not up-to-date |  | 11.1% (7) | 24.4% (253) |
|  | Missing data |  | 73.0% (46) | 35.6% (368) |

<sup>1</sup>Data reported as % (n) and mean ± SD
<sup>2</sup>Calculated only for those children without oedema
<sup>3</sup>Based on children ≥12 months who had received all vaccinations per ministry of health, Ugandan guidelines including measles vaccine (9 months), DPT3 (14 weeks), vitamin A (at 6 months), and deworming (≥12 months)

Although all children admitted at the unit were diagnosed with cSAM, after re-analysis, a few of those diagnosed with complicated non-oedematous SAM actually had MAM (3.2% vs 6.2% among relapses vs non-relapses respectively). At their index admission, children who relapsed weighed slightly more (6.8 ± 2.1 kg) than non-relapses (6.0 ± 2.9 kg). Both groups nearly had same MUAC (∼11.0 cm), however, relapses were slightly more wasted (WHZ: -4.4 ± 1.1) than non-relapses (-3.8 ± 1.6) (Table 2). A higher proportion of relapsed children had missing data especially regarding their immunization status (73.0%).

After multiple logistic regression, older children had 0.05 (95%CI: 0.02; 0.08) higher odds of relapsing with cSAM compared to their younger counterparts (**Table 3**). Additionally, for every one more surviving sibling at home, there was a 0.3 (95%CI: 0.09; 0.51) higher odd of relapsing with cSAM. Compared to children whose mothers were in some formal/informal employment, those with unemployed mothers were associated with 2.63 (95%CI: 0.61; 4.65) higher odds of relapsing with cSAM (Table 3). There was no observed association between cSAM relapse with either any nutritional or clinical factor.

**Table 3:** Socioeconomic, nutritional, and paraclinical correlates of complicated severe acute malnutrition relapse among 1098 children admitted at Mwanamugimu^1^.

| Characteristics | OR (95% CI) | p |
| --- | --- | --- |
| **Sociodemographic and socioeconomic status** |  |  |
| Age | 0.05 (0.02; 0.08) | **0.004** |
| Sex (male) | -0.48 (-1.37; 0.41) | 0.29 |
| Residence Kampala | Ref | Ref |
| Wakiso | -0.22 (-1.16; 0.71) | 0.64 |
| Other districts | -0.09 (-1.11; 0.94) | 0.87 |
| Caregiver (mother) | -0.13 (-1.23; 0.97) | 0.82 |
| Number of surviving children at home | 0.30 (0.09; 0.51) | **0.01** |
| Caregiver's marital status (married) | -0.23 (-1.05; 0.59) | 0.59 |
| Maternal occupation |  |  |
| Employed | Ref | Ref |
| Unemployed | 2.63 (0.61; 4.65) | **0.01** |
| Missing data | 1.86 (-0.30; 4.02) | 0.09 |
| **Nutritional status** |  |  |
| Nutrition diagnosis |  |  |
| Severe acute malnutrition non-oedematous | Ref | Ref |
| Severe acute malnutrition oedematous | -0.28 (-1.14; 0.58) | 0.53 |
| Moderate acute malnutrition | 0.21 (-1.33; 1.75) | 0.79 |
| Weight (kg) | -0.08 (-0.32; 0.17) | 0.54 |
| Mid-upper arm circumference (cm) | 0.54 (-0.23; 1.32) | 0.17 |
| Weight-for-age z-score | 0.09 (-0.29; 0.47) | 0.63 |
| Height-for-age z-score | 0.03 (-0.20; 0.25) | 0.82 |
| Weight-for-height z-score<sup>2</sup> | -0.27 (-0.61; 0.07) | 0.13 |
| **Paraclinical status** |  |  |
| HIV status |  |  |
| Seropositive | Ref | Ref |
| Seronegative | -0.54 (-1.51; 0.44) | 0.28 |
| Unknown | 0.21 (-0.92; 1.35) | 0.71 |
| Missing data | 0.23 (-1.93; 2.39) | 0.84 |
| Immunization |  |  |
| Up-to-date<sup>3</sup> | Ref | Ref |
| Not up-to-date | 0.63 (-0.40; 1.67) | 0.23 |
| Missing data | 0.28 (-0.69; 1.24) | 0.57 |

<sup>1</sup>Data reported as logistic regression odds ratio, OR (95% CI), adjusted for age at index admission, and sex
<sup>2</sup>Calculated only for those children without oedema
<sup>3</sup>Based on children who had received all vaccinations (for age) per ministry of health, Ugandan guidelines

## Discussion

Our findings reveal that about 6% of children under 5 are at risk of relapsing with cSAM at Mwanamugimu, Mulago hospital. Due to lack of a definition and set cSAM threshold in either the national guidelines [8] or international standards [10], our prevalence can neither be deemed acceptable nor unacceptable. Factors associated with cSAM relapse included older age, living with more siblings at home, and those whose mothers were unemployed. Although few studies have looked at relapse with cSAM, a similar study in Zambia and Zimbabwe found all-cause hospital readmission of 15.4% 52-weeks post-discharge [12]. In their study, 52.8% of readmissions occurred in the first 12 weeks post discharge. In a review of relapse following recovery from SAM treatment, the proportions varied from 0 – 37% across varied lengths of time up to 18-months post-discharge [9] with the highest proportions noted within 6-months. This systemic review did not necessarily look at cSAM but rather all SAM cases managed in the CMAM programs. The proportional range from this systemic review also encompasses the 17.8% relapse rate reported earlier in a 6-months follow-up CMAM program in Bangladesh [13]. This follow-up study enrolled younger children (6-23 mo) having SAM with acute illness that attended a day care clinic where they received antimicrobials, micronutrients, and milk-based diets until clinical improvement [13]. Thereafter, they were transitioned to a day-care nutrition rehabilitation unit receiving only nutritious diets until attainment of 80% WHZ and would be considered relapses after experiencing a new SAM episode. Our proportion of cSAM relapse (5.7%) is largely incomparable across studies due to various reasons but in view of the elaborate management criteria at such units like MNU, almost none of the children would be expected to be readmitted post-discharge. Moreover, all admitted children are investigated for any underlying illnesses, started on appropriate care if chronic, caregivers counseled on age-appropriate IYCF with practicum, and linked to community livelihood and other socioeconomic transformation programs at discharge.

The association between age and child undernutrition has been established in various literature [14, 15]. It is noted that as children grow from infancy, they are gradually weaned from breastmilk onto the staple mainly comprising of starchy cereal gruel [16]. Due to such poor complementary feeding practices, injustice is done early especially regarding children’s nutritional requirements vis-à-vis what’s supplied likely culminating into undernutrition. To add to this predicament, with increased number of young children, more attention is given to the younger one often neglecting the older ones. Our findings similar to other studies [17, 18] show that increased family size was associated with cSAM relapse. This is also premised on the assumption that with increased number of children, there is an increased burden on family resources including food consumption putting them at higher risk of food insecurity and poverty [18]. These tend to switch to such coping mechanisms like consuming less quantity, fewer times, moreover without minding the quality of food.

Using maternal unemployment as a proxy of poor household income, our results agree with existing data regarding the established association with child undernutrition [18, 19]. Households with unemployed mothers are likely to have challenges in accessing enough quantity of nutritious food ultimately leading to food and nutrition insecurity [18]. This not only affects the dietary intake of children but also the care practices in poor homes are sub optimal affecting several child rearing activities including access to health services and stimulation.

Key strength of our study is we had a large sample size of cSAM children enabling us to examine some associations with good power. However, among other weaknesses, using secondary data posed a challenge of missing data moreover inconsistent across variables. Additionally, we were unable to obtain a good measure of such variables like household socioeconomic status but used appropriate proxy. There was no standard definition of the outcome ‘cSAM relapse’ and, were incapable of comparing across several studies partly due to difference in treatment protocols, varied time-to-relapse used, and reports of relapse (point prevalence/cumulative/incident rate) as admissible elsewhere [9]. We were unable to estimate the actual risk attributable to relapsing with cSAM at MNU, Mulago hospital given our study type.

### Conclusion and recommendations

Our findings reveal an unignorable burden of cases of relapse with complicated SAM in inpatient therapeutic care centers with some often going unnoticed. Together with the established associated factors, we add to the call to have a standardized definition of cSAM relapse, consideration for its inclusion as one of the performance indicators with a set threshold.

For ITC forefront medical personnel, attention should be taken at admission to establish those being readmitted. The centers may also need to reinforce their community linkages to ensure that poor and large households are prioritized for livelihood support among other underlying factors, to alleviate risk of relapses in such LMICs.

Later, perhaps with a standard definition, a longitudinal follow-up study of children discharged from ITCs while assessing key potential risk factors may be justified to establish who is most at risk of relapsing and why.

## Data Availability

The Ugandan act on Data Protection does not allow for personal data to be made available to other researchers without prior written approval from relevant institutions and authorities. In case of any queries, the Corresponding author can be contacted at

## Author contributions

JM conceptualized the study; JM developed the proposal; JM oversaw the data collection, entry, and cleaning; The statistical analysis plan was developed by JM and Victoria Nankabirwa and analysis done by JM with both interpreting the results; This manuscript was written by JM; JM took final responsibility on the publication plan.

